# Altered placental immune cell composition and gene expression with isolated fetal spina bifida

**DOI:** 10.1101/2023.11.24.23298970

**Authors:** Marina White, Hasan Abdo, David Grynspan, Tim Van Mieghem, Kristin L Connor

**Affiliations:** Department of Health Sciences, Carleton University, Ottawa, ON K1S 5B6, Canada; Vernon Jubilee Hospital, Vernon, BC V1T 5L2, Canada; Department of Pathology and Laboratory Medicine, University of British Columbia, Vancouver, BC V6T 1Z7, Canada; Department of Obstetrics and Gynecology, Sinai Health System, Toronto, ON M5G 1X5, Canada

**Author notes:** Correspondence: Dr. Kristin Connor, Department of Health Sciences, Carleton University, Ottawa, Ontario, Canada, K1S5B6, E.

**Keywords:** Neural tube defects, spina bifida, placenta, folate, Hofbauer cells

## Abstract

**Problem:** Maternal B vitamin deficiency increases the risk of fetal spina bifida (SB) and placental maldevelopment. It is unclear whether placental processes involving folate are altered in fetuses with SB in a contemporary cohort. We hypothesised that fetal SB would associate with reduced expression of key folate transporters (folate receptor-α [FRα], proton coupled folate receptor [PCFT], and reduced folate carrier [RFC]), and an increase in Hofbauer cell (HBC) abundance and folate receptor-*β* (*FRβ)* expression by HBCs.

**Method of Study:** FRα, PCFT, and RFC protein localisation and expression (immunohistochemistry) and HBC phenotypes (RNA *in situ* hybridization) were assessed in placentae from fetuses with SB (cases; n=12) and with no congenital anomalies (controls; n=22).

**Results:** Cases (vs. gestational age [GA]-matched controls) had a higher proportion of placental villous cells that were HBCs (6.9% vs. 2.4%, p=0.0001) and higher average *FRβ* expression by HBCs (3.2 mRNA molecules per HBC vs. 2.3, p=0.03). HBCs in cases were largely polarised to a regulatory phenotype (median 92.1% of HBCs). In sex-stratified analyses, male, but not female, cases had higher HBC levels and *FRβ* expression by HBCs than GA-matched controls. There were no differences between groups in the total percent of syncytium and stromal cells that were positive for FRα, PCFT, or RFC protein immunolabelling.

**Conclusions:** HBC abundance and *FRβ* expression by HBCs are increased in placentae of fetuses with isolated SB, suggesting immune-mediated dysregulation in placental development and function, and could contribute to SB-associated comorbidities, such as poor fetal growth.

## Introduction

Maternal periconceptional deficiencies of folate and folate metabolism co-factors are known to increase the risk of fetal neural tube defects (NTDs)^1-3^. Around half of NTDs are estimated to be responsive to folates^3^, which are essential, water-soluble B vitamins required for the high rates of DNA synthesis and cell proliferation that accompany periods of rapid development (including neural tube closure)^4^. However, folates are essential throughout pregnancy, well-beyond the period of neural tube closure, to support fetoplacental growth and placental function^5^. It is unclear whether placentae from fetuses with NTDs may be at increased risk of dysfunction in processes involving folate, and whether these disruptions may contribute to the poor growth^6,7^ and early birth^8,9^ experienced by these fetuses^10-12^.

Hofbauer cells (HBCs) are placental macrophages of fetal origin that highly express folate receptor-β (FRβ), one of the major placental folate transporters^13-15^. HBCs favour polarisation towards a regulatory (M2) phenotype^16^, and as a result, play a critical role in the regulation of placental angiogenesis, immune function, and inflammation^15^. Although HBCs are detectable as early as 18 days post-fertilization^14,15^, HBC ontogeny across gestation is not well defined. Importantly, as HBCs express *FRβ*, it is possible that beginning early in pregnancy, HBC level and function could be sensitive to changes in local folate bioavailability and/or metabolism. In assessments of placental histopathology from both historical (pre-folic acid fortification/supplementation) and contemporary cohorts^12,17^, we have shown that fetuses with NTDs have increased HBC abundance. However, how HBC phenotype may be altered in fetuses with NTDs remains unknown. It is plausible that a uterine environment permissive of NTDs may influence HBC phenotype and function, and that these changes could contribute to placental villous maldevelopment, as observed in these cases^12,17^.

Folates are also actively transported to the fetus to support fetal growth and development through transmembrane transporters^18^. Folate uptake and transport across the placental syncytium is coordinated by folate receptor-α (FRα), proton coupled folate receptor (PCFT/heme carrier protein 1 [HCP1]), and reduced folate carrier (RFC)^14,18,19^. Reduced activity in these transporters is associated with suboptimal fetal outcomes, such as intrauterine growth restriction^20^. Conversely, in fetuses with NTDs, an adaptive upregulation of placental *FRα* and *RFC* expression has been observed in response to low folate bioavailability^21^. However, it remains unclear if placental folate transport in fetuses with NTDs may be altered in regions with a low burden of folate deficiency, where maternal folate levels are likely adequate, and whether compromised placental folate transport may relate to the poor growth outcomes often experienced by these fetuses^10-12^.

Here we aimed to better understand whether, and how, HBC phenotype and placental folate transport were altered in fetuses with isolated spina bifida (SB; the most common NTD) compared to placentae from fetuses with no congenital anomalies. We also aimed to understand whether changes in placental HBC phenotype and folate transport differed based on placental sex. We hypothesised that cases would have a higher number of HBCs, and that HBCs would have increased *FRβ* expression and adopt a more activated (*CD80*^*+*^) phenotype. We also hypothesised that cases would have reduced expression of folate transporters, increasing the risk of poor fetal growth. Lastly, to understand the mechanisms that may underly changes in placental phenotype, we explored relationships between key maternal and infant characteristics, HBC phenotype, and placental folate transport.

## Methods

### Ethics

This study was approved by the Carleton University Research Ethics Board (108884 and 106932) and the Mount Sinai Hospital Research Ethics Board (17-0028-E and 17-0186-E).

### Population and study design

This study population has been previously described^22^. Pregnant people carrying a fetus were recruited at approximately 25 weeks’ gestation at Mount Sinai Hospital, Toronto. Controls were singleton pregnancies with no known fetal anomalies. At recruitment, maternal clinical data, medical history, and dietary recall questionnaire data were collected, and data on pregnancy and birth outcomes were collected at delivery. A second control group matched to cases for gestational age (GA) at delivery was also identified (preterm controls [hereon referred to as PT controls]) at the same site. PT controls were singleton pregnancies with no known fetal anomalies or maternal pregnancy complications^22^, and were matched 1:1 with cases for GA at delivery and maternal pre-pregnancy body mass index (BMI)^23^.

### Maternal clinical, demographic, and dietary characteristics

Maternal clinical data and medical history, and dietary recall questionnaire data (Automated Self-Administered 24-hour Recall™, specific to the Canadian population [ASA24-Canada-2016]^24^) were collected at recruitment for cases and controls. The maternal dietary recall data have been previously reported in full^22^, and here, we analysed reported maternal intakes of folate and vitamins B6 and B12 as exposure variables of interest.

### Infant birth outcomes

Data on pregnancy and infant birth outcomes have been previously reported for this cohort^22^. GA at delivery was calculated using maternal reports of last menstrual period to the nearest week. Birthweight z-scores adjusted for infant GA and sex were calculated using Canadian growth standards^25^.

### Placental collection and processing

#### Sample collection

All placental samples were collected and processed by the Research Centre for Women’s and Infants’ Health BioBank immediately after delivery. Placental biopsies were collected from each of the four placental quadrants, at least 1.5 cm away from the centre and edge of the placental disk, fixed in paraformaldehyde and embedded in paraffin.

#### Immunohistochemistry

Immunohistochemistry (IHC) was used to assess placental folate transporter localisation and immunoreactive (ir) protein expression. Paraformaldehyde-fixed and paraffin-embedded (PFPE) placental samples were cut at 5 μm and placed on a slide warmer (Agar Scientific, Stansted, United Kingdom) for 30 minutes at 55°C. Placental sections were stained using rabbit polyclonal antibodies to visualize FRα (Anti-Folate Binding Protein/FBP; ab67422; 1:250), PCFT (Anti-HCP1/PCFT; Abcam ab25134; 1:100), and RFC (Anti-RFC; Abcam ab62302; 1:150). The slides were processed using a protocol previously described^26^, with a few modifications. In brief, after slides were deparaffinized in xylene and rehydrated in ethanol dilutions, a heat-induced antigen retrieval was performed in a microwave using a 10mM citric acid and sodium citrate (pH 6.0) solution. Slides were then washed with 1X phosphate-buffered saline (PBS), quenched using 3.0% hydrogen peroxide in 97% methanol to block endogenous peroxidase activity, and incubated with a protein-blocking solution (Agilent Dako, Santa Clara, USA) for one hour at room temperature to block non-specific binding. Slides were incubated overnight with each specific primary antibody at 4°C. A negative control was included in each run, for which the primary antibody was replaced with antibody diluent. Slides were incubated with a secondary antibody (1:200 goat biotinylated anti-rabbit antibody, BA-1000; Vector Labs, Burlingame, USA) and then streptavidin-horse radish peroxidase (1:2000 dilution in 1X PBS; Invitrogen, Carlsbad, USA) for one hour each at room temperature. Horse radish peroxidase enzymatic activity was detected through treatment with 3,3’ diaminobenzidine (DAB; Vectastain Elite ABC HRP Kit, Vector Laboratories, Brockville, Canada) for two minutes. Gill’s #1 haematoxylin (Sigma-Aldrich, Oakville, Canada) was used to counterstain the slides.

Whole placental sections from IHC analyses were scanned on the EVOS FL Auto 2 at 20X using the automate function (Thermo Fisher Scientific, Waltham, MA, USA). The boundaries of the placental villous tissue on each slide were manually indicated within the software, and after scanning, unstitched image files were exported (range: 207-961 fields, depending on the size of placental tissue). A file list, where each file name corresponds to an image (field) captured, was generated for each antibody for each placenta. From this file list, 50 images from each placental sample were randomly selected for analyses using the ‘=rand()’ and ‘=index()’ functions in Excel.

Localisation and semi-quantitative expression of immunoreactive FRα, PCFT and RFC was assessed using QuPath (version 0.3.2)^27^, which has previously been used to analyse IHC images in placental tissue^28,29^. For each sample, colour deconvolution was performed (‘estimate stain vectors’ tool), the intensity of staining was evaluated (‘positive cell detection’ tool^28^), and the detection parameters were optimised (Supplementary Table S1 and Figure S1). The positive cells were classified as stained with low (threshold 1), medium (threshold 2), or high (threshold 3) intensity in alignment with visual assessments (Supplementary Table S1 and Figure S1). Percent syncytium and stromal cells stained overall, and at low, medium, and high thresholds, was quantified for each antibody.

#### RNA in situ hybridization

To evaluate the abundance, polarisation, and cellular origins of HBCs, as well as *FRβ* expression by HBCs, *in situ* hybridization (ISH) was performed using the RNAscope^™^ Multiplex Fluorescent v2 Assay (Cat. No. 323100; ACD-Bio). Following the manufacturer’s protocol^30,31^, PFPE placental samples were cut at 5 μm and placed on a slide warmer (Agar Scientific, Stansted, United Kingdom) for 30 minutes at 55°C. Following de-paraffinization, target retrieval and protease treatment^30,31^, the slides were treated with probes specific to *FRβ* (pan-HBC marker^14^; channel 1), vimentin (*VIM*; marker for cells of mesenchymal origin^32^; channel 2), cluster of differentiation 80 (*CD80*; activated HBC marker^33^; channel 3), and mannose receptor C-type 1 (*MRC1/CD206*; regulatory HBC marker^33-35^; channel 4). Slides treated with ready-to-use negative control (*dapB*) and positive control probes (four probes targeting DNA-directed RNA polymerase II subunit RPB1 [*POLR2A*], peptidyl-prolyl cis-trans isomerase B [*PPIB*], ubiquitin C [*UBC*], and hypoxanthine phosphoribosyltransferase 1 [*HPRT-1*]) were also run in each batch. All probes were tested on known positive control tissues (RNAscope® Human Hela Cell pellet; Cat No. 310045) prior to use on human placental tissues. Following the amplification steps, the target probes were labelled using Opal dyes (Opal 520 [FP1487001KT; labelled *MRC1*], Opal 570 [FP1488001KT; labelled *VIM*], Opal 620 [FP1495001KT; labelled *FRβ*], and Opal 690 [FP1497001KT; labelled *CD80*]; Akoya Biosciences). The slides were counterstained using DAPI (ACD-Bio)^30,31^. Slides treated with a single Opal fluorophore for all channels were also run in each batch, to use as reference spectra a later stage for spectral unmixing.

Whole placental sections from ISH analyses were captured as Z-stack images at 20X on the Axio Scan Z1 Slide Scanner (ZEISS, Oberkochen, Germany). Five fluorescent channels were used simultaneously: Alexa Fluor (AF) 488 (Opal 520/*MRC1*/*CD206* probe), AF568 (Opal 570/*VIM* probe), AF594 (Opal 620/*FRβ* probe), AF647 (Opal 690/*CD80* probe), and a 405nm diode laser for DAPI. ZEN lite (version 3.4) was used to process the images (CZI files). Following the calculation of average intensity projections for each pixel (orthogonal projection), linear unmixing was performed using the reference spectra for each Opal fluorophore, and reference spectra for DAPI obtained from a negative control slide. High levels of red blood cell autofluorescence in the green channel (AF488) rendered any probe signal in this channel undetectable, and this channel (tagged with the *MRC1*/*CD206* probe) was excluded from subsequent analyses.

Linearly unmixed images (as CZI files) were exported for subsequent analysis in QuPath (version 0.3.2)^27^, to assess the localisation and expression of target probes. Unmixed CZI files were imported into QuPath (Bio-Formats). Guided by the assay manufacturer’s technical note on using QuPath to analyze RNAScope^™^ images^35^, cell detection parameters (to detect DAPI signal) were optimized from the default settings by adjusting: the setup parameters (requested pixel size [to improve image processing resolution]), intensity parameters (threshold [to maximize nuclei detection, including faintly stained nuclei]), and cell parameters (cell expansion [to define the cell area that surrounds the cell nucleus). The cell boundaries were defined based on known HBC size (10–30 μm^31^), such that cells were defined as the 10 μm that surrounded the nucleus on any side, while taking into consideration the presence of other nearby cells. No probe was counted twice (i.e., each signal could only be counted once and in relation to a single cell; an illustration of this is provided in Supplementary Figure S2). To identify cells that expressed the target probes, single measurement classifiers were used to detect each of the target probe’s signals, within each of the cell’s boundaries.

The RNAScope^™^ signal is displayed as punctate dots, where each dot represents a single copy of an mRNA molecule for the target gene and larger dots or clusters may indicate the presence of multiple mRNA molecules in proximity. A cell tagged with one or more punctate dots was classified as ‘positive’ for a given target probe. Cells were classified as: *FRβ*^*+*^ (HBCs), *CD80*^*+*^ (cells expressing *CD80*), *VIM*^*+*^ (cells of mesenchymal origin). Composite cell classifiers were then applied to identify cells that were positive for more than one probe, including cells that were: *FRβ*^*+*^/*CD80*^*+*^ (Activated/ *CD80*^+^ polarised HBCs), *FRβ*^*+*^/*VIM*^*+*^ (HBCs of mesenchymal origin), or *FRβ*^*+*^/*CD80*^*+*^/*VIM*^*+*^ (Activated/ *CD80*^+^ polarised HBCs of mesenchymal origin). The number of detections for each classifier was summed across 20 fields of view for each placental sample to obtain a total detection count for each classifier per sample. Then, for each sample, these classifiers were used to calculate the following:

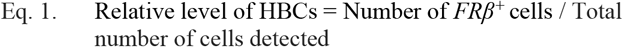

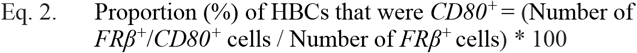

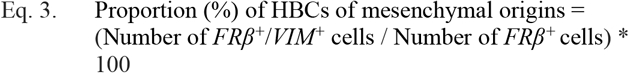

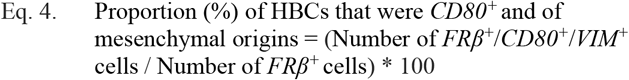

From these proportions (equations 2-4), we deduced the proportion of HBCs that were *CD80*^*−*^ (100 – proportion [%] of HBCs that were *CD80*^*+*^), of non-mesenchymal origins (100 – proportion [%] of HBCs of mesenchymal origins), or both (100 – proportion [%] of HBCs that were *CD80*^*+*^ and of mesenchymal origins).

We also used the sub-cellular detection classifier in QuPath to quantify the level of *FRβ* expression by HBCs for each sample (i.e., the number of *FRβ* dots on each *FRβ*^*+*^ cell detected across 20 fields of view), which was used to determine:

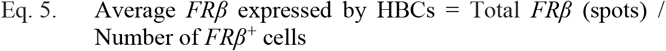

A script was generated and applied to process all annotated fields of view, for each sample, using the optimised analysis settings and the batch processing function in QuPath (Supplementary Table S2 and Figure S2A-D).

### Statistical analysis

Data were analysed using JMP (version 16.0, SAS Institute Inc, Cary, NC). Associations between study group with: 1. percent syncytium and stromal cells stained with FRα, PCFT and RFC (overall, and at low, medium, and high thresholds), and 2. HBC phenotype variables (relative level, polarisation, origins and average *FRβ* expression) were tested using unadjusted (ANOVA, for parametric data; Wilcoxon test, for non-parametric data) and adjusted (standard least squares regression and Tukey’s post hoc test) models. All adjusted models included infant sex (male, female; categorical) and gestational age at delivery (20-41 weeks; continuous). Data are reported as median (IQR) with p values from standard least squares regression (adjusted). Statistical significance was set at p<0.05.

To determine whether key maternal and infant characteristics may relate to placental folate transporter/receptor expression or HBC phenotype, we also explored associations between maternal pre-pregnancy BMI and reported B vitamin intakes, and infant GA at delivery with HBC phenotype and placental folate transport, using Spearman’s rank correlation (rho [ρ]) test. Statistical significance was set at p<0.05.

## Results

### Clinical cohort characteristics

Baseline characteristics for PT controls (n=12), controls (n=10), and cases (n=12) have been previously described and the groups were highly similar^22^. This is likely a folate replete population, given that all mothers in the case and control groups reported taking prenatal vitamins and that in Canada, fortification of white flour, enriched pasta and corn meal with folic acid is mandatory^22^.

### Placental folate transporter expression did not differ between cases and controls

FRα protein immunolabelling was largely localized to the syncytiotrophoblast, with some staining in villous core connective tissue cells and the endothelial cells of the fetal capillaries (Figure 1). A high degree of PCFT and RFC protein immunolabelling was visible in the syncytiotrophoblast and cytotrophoblast cells layers, as well as in the villous core connective tissue cells and the endothelial cells of the fetal capillaries (Figure 1). There were no differences between cases, controls and PT controls in the total percent of syncytium and stromal cells that were positive for FRα, PCFT, or RFC protein immunolabeling (Figure 2A-B, Supplementary Table S3). There were also no differences between cases, controls, and PT controls in the percent of syncytium and stromal cells that were positive for FRα, PCFT, or RFC protein immunolabeling at the low, medium, and high staining intensity thresholds (Figure 2A-B, Supplementary Table S3).

**Figure 1.**
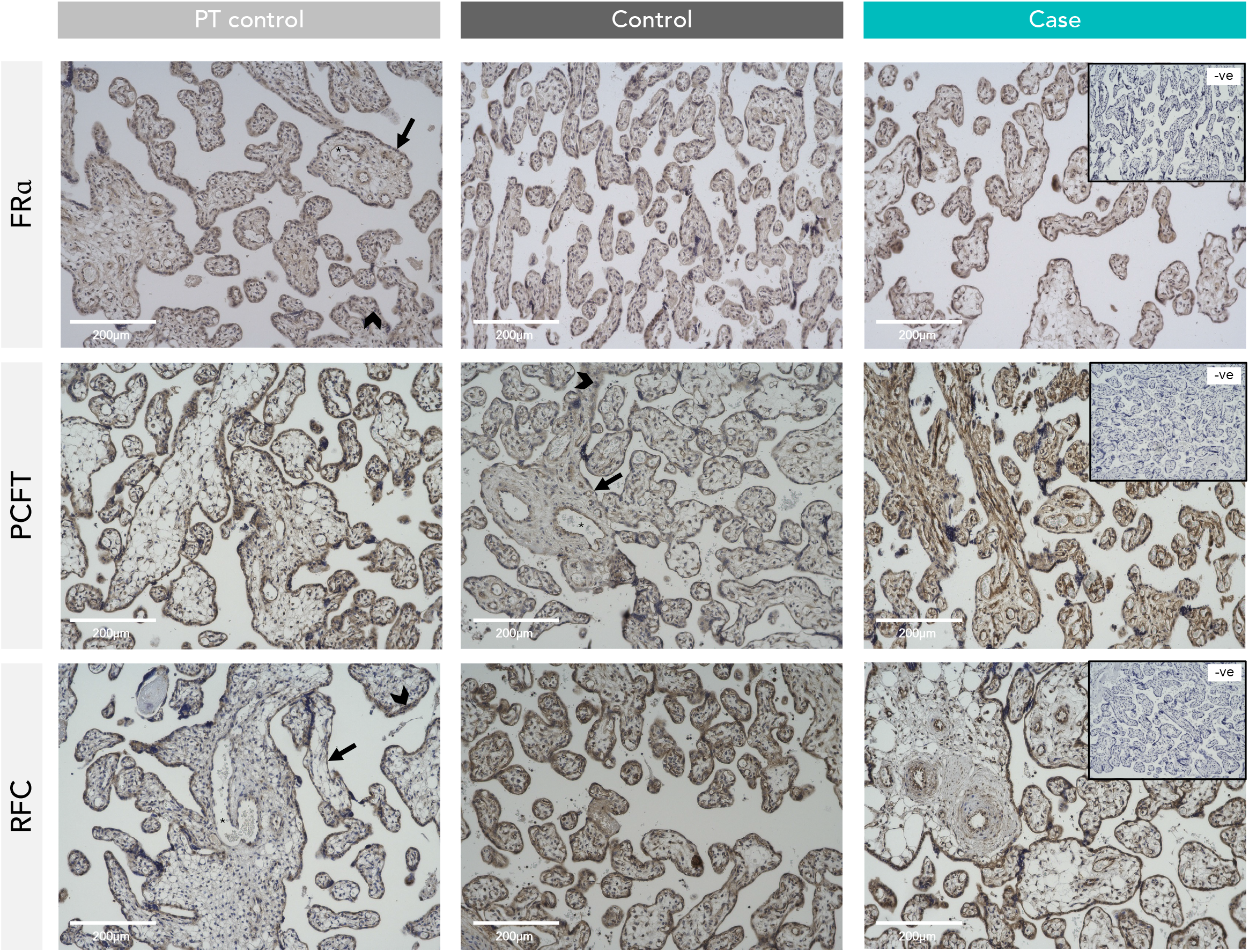
Representative IHC images of FRα, PCFT, and RFC protein staining in human placenta across study groups. Arrow = syncytiotrophoblast. Arrowhead = cytotrophoblast. Asterisk = fetal blood vessels. FRα = folate receptor alpha. PCFT = proton coupled folate transporter. RFC = reduced folate carrier. PT-controls (n=12), controls (n=10), and cases (n=12).

**Figure 2.**
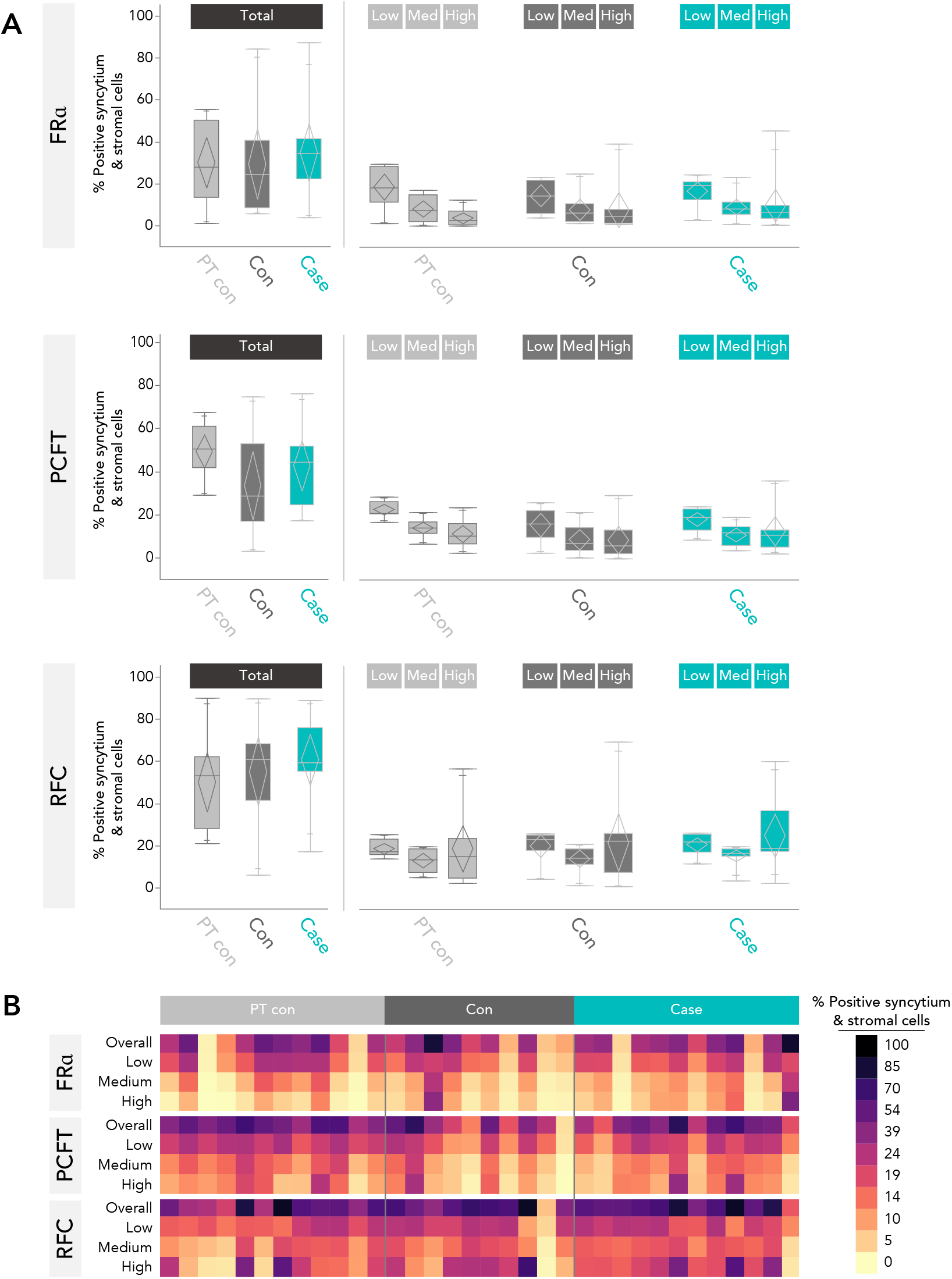
FRα, PCFT, and RFC protein staining of immunolabeled areas in placental syncytium and stromal cells. (A) Boxplots of FRα, PCFT, and RFC protein staining of immunolabeled areas in placental syncytium and stromal cells in PT-controls (n=12) and controls (n=10), and cases (n=12). There were no differences for total percent FRα-, PCFT-, or RFC-positive syncytium and stromal cells, or percent FRα-, PCFT- and RFC-positive syncytium and stromal cells at the low, medium, and high staining intensity thresholds, between cases, controls and PT controls. (B) Heatmap of total and low, medium, and high intensity staining of immunolabeled areas in placental syncytium and stromal cells. Data presented are from a single experiment. Data from semi-quantitative analysis of staining in immunolabeled areas were analyzed using standard least squares regression models, adjusted for infant sex and gestational age at delivery. FRα = folate receptor alpha. PCFT = proton coupled folate transporter. RFC = reduced folate carrier.

### HBC abundance and FRβ expression were increased in cases compared to PT controls

Cases had a higher proportion of placental villous cells that were HBCs (*FRβ*^+^ cells) than GA-matched PT controls (6.9% [4.3, 10.2] vs. 2.4% [1.2, 3.2], p=0.0001; Figure 3B-C, Supplementary Table S4). Average *FRβ* expression by HBCs, determined by the number of *FRβ* spots (mRNA molecules) per HBC, was also higher in cases than PT controls (3.2 mRNA molecules per HBC [2.5, 4.2] vs. 2.3 [2.1, 2.6], p=0.03; Figure 3D, Supplementary Table S4). There were no differences between cases and controls for the proportion of placental villous cells that were HBCs, or average *FRβ* expression by HBCs (Figure 3B-D, Supplementary Table S4).

**Figure 3.**
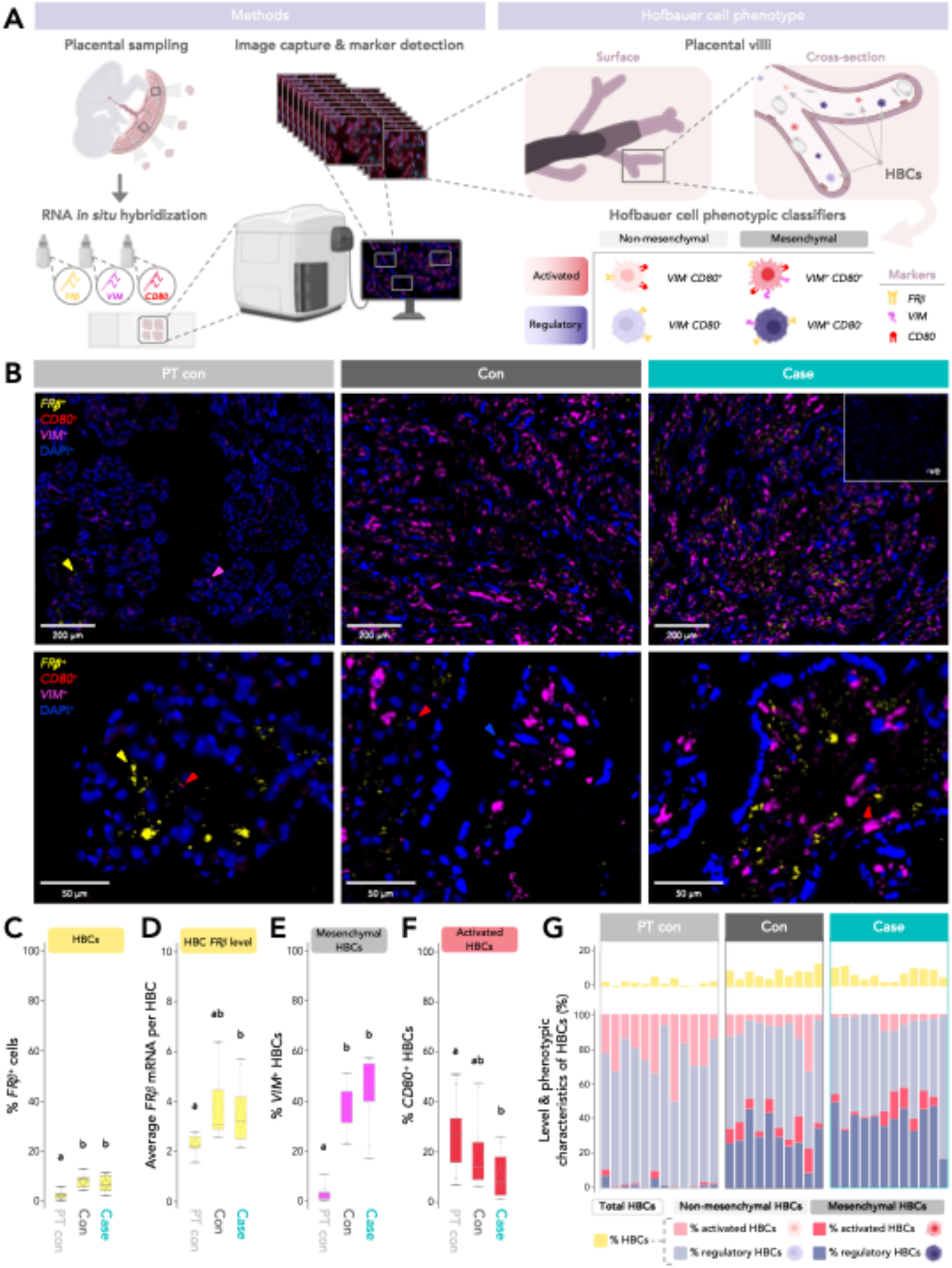
Hofbauer cell detection and phenotype in cases, PT-controls, and controls. (A) Visual schematic of the methods used to profile HBC phenotype in human placentae, and the HBC phenotypic classifiers that were considered. (B) Representative images from RNA *in situ* hybridization using target probes specific to human *FRβ* (yellow), *VIM* (pink), *and CD80* (red), and DAPI nuclear stain (blue) on human placental villous tissue. Top panels left-to-right show probe detection in PT controls (n=12), controls (n=10), and cases (n=12). Bottom panels show a closer frame from the same samples pictured in the top panels. One dot = one mRNA molecule. Triangles = probe staining (for each respective colour). Arrowhead = syncytiotrophoblast. Cases had a higher proportion of (C) % *FRβ*+ cells (HBCs; pan-HBC marker), (D) average *FRβ* expression by HBCs, and (E) *VIM*^*+*^ HBCs (HBCs with mesenchymal origins) than PT controls. (F) Cases had a lower proportion of *CD80*^*+*^ HBCs than PT controls. (G) A stacked bar chart visualising the distribution of HBC phenotypes observed (% HBCs with mesenchymal and non-mesenchymal origins, polarised to activated or regulatory phenotypes). Pink = activated HBCs. Purple = regulatory HBCs. Bright colours = HBCs with mesenchymal origins. Faded colours = HBCs with non-mesenchymal origins. Yellow = the proportion (%) of total cells that were HBCs. Data presented are from a single experiment. Data were analysed using standard least squares regression models, adjusted for infant sex and gestational age at delivery. HBC = Hofbauer cell. FRβ = folate receptor beta. CD80 = cluster of differentiation 80. VIM = vimentin.

### A higher proportion of HBCs in cases were of mesenchymal origin and CD80^-^ compared to PT controls

The proportion of HBCs that were of mesenchymal origins (*VIM*^*+*^) was higher in cases than PT controls (43.7% [39.8, 54.9 vs. 1.8% [0.7, 3.3], p<0.0001; Figure 3E, Supplementary Table S4). Overall, the proportion of HBCs with mesenchymal origins in PT controls was low, indicating a dominance of HBCs of non-mesenchymal origins (Figure 3B and E). There were no differences between cases and controls for the proportion of HBCs that were of mesenchymal origins (Figure 3E, Supplementary Table S4).

Cases had a lower proportion of HBCs that were polarised to an activated phenotype (*CD80*^*+*^) than PT-controls overall (7.9% [2.6, 17.7] vs. 26.6% [15.8, 33.3], p=0.01; Figure 3F, Supplementary Table S4). Cases also had a lower proportion of activated (*CD80*^*+*^) HBCs with non-mesenchymal origins (*VIM*^−^) than PT-controls (3% [1.9, 6.3] vs. 24.8% [15.2, 32.5], p=0.00003), but a higher proportion of activated HBCs that were of mesenchymal origins (*VIM*^+^) than PT-controls (5% [0.9, 10] vs. 0.61% [0.22, 1.3], p=0.02; Supplementary Table S4). Conversely, the proportion of HBCs that were *CD80*^−^ was higher in cases than PT-controls in HBCs of both mesenchymal and non-mesenchymal origins (Supplementary Table S4). HBC origins (mesenchymal and non-mesenchymal) did not appear to influence likelihood of HBC polarisation to an activated phenotype in cases (Supplementary Table S4). There were no differences between cases and controls in HBC polarisation, overall or within HBC subgroups stratified by mesenchymal and non-mesenchymal origins (Figure 3F, Supplementary Table S4).

HBC phenotype distribution (mesenchymal and non-mesenchymal origins, polarised to activated or regulatory phenotypes) is visualised in Figure 3G. *CD80*^−^ HBCs comprised the majority of HBCs for all samples (except one PT control), and five cases had a low total proportion (<4%) of HBCs that were polarised to an activated phenotype (Figure 3G).

### HBC phenotype in cases may differ by fetal sex

In exploratory analyses of folate-sensitive placental outcomes stratified by fetal sex, we found that male cases had a higher proportion of placental villous cells that were HBCs (*FRβ*^+^ cells) than male GA-matched PT controls (10% [7.8, 11.3] vs. 2.4% [1.8, 4.5], p=0.002; Table 1). Male cases also had higher average *FRβ* expression by HBCs (3.6 *FRβ* mRNA molecules per HBC [2.9, 4.6] vs. 2.3 [2.1, 2.6], p=0.01), and a higher proportion of HBCs with mesenchymal origins (*VIM*^*+*^; 43.7% [39.8, 54.9 vs. 1.8% [0.7, 3.3], p<0.0001) than male PT controls (Table 1). There were no differences in the proportion of HBCs that were activated (*CD80*^*+*^), or FRα, PCFT, and RFC protein immunolabeling in male cases compared to male PT controls, and no differences in HBC phenotype or placental folate transporter protein immunolabeling in male cases compared to male controls (Table 1). However, it should be noted that as only a small proportion of cases (4 of 12) and controls (2 of 10) were male, these findings should be interpreted with caution.

**Table 1.**
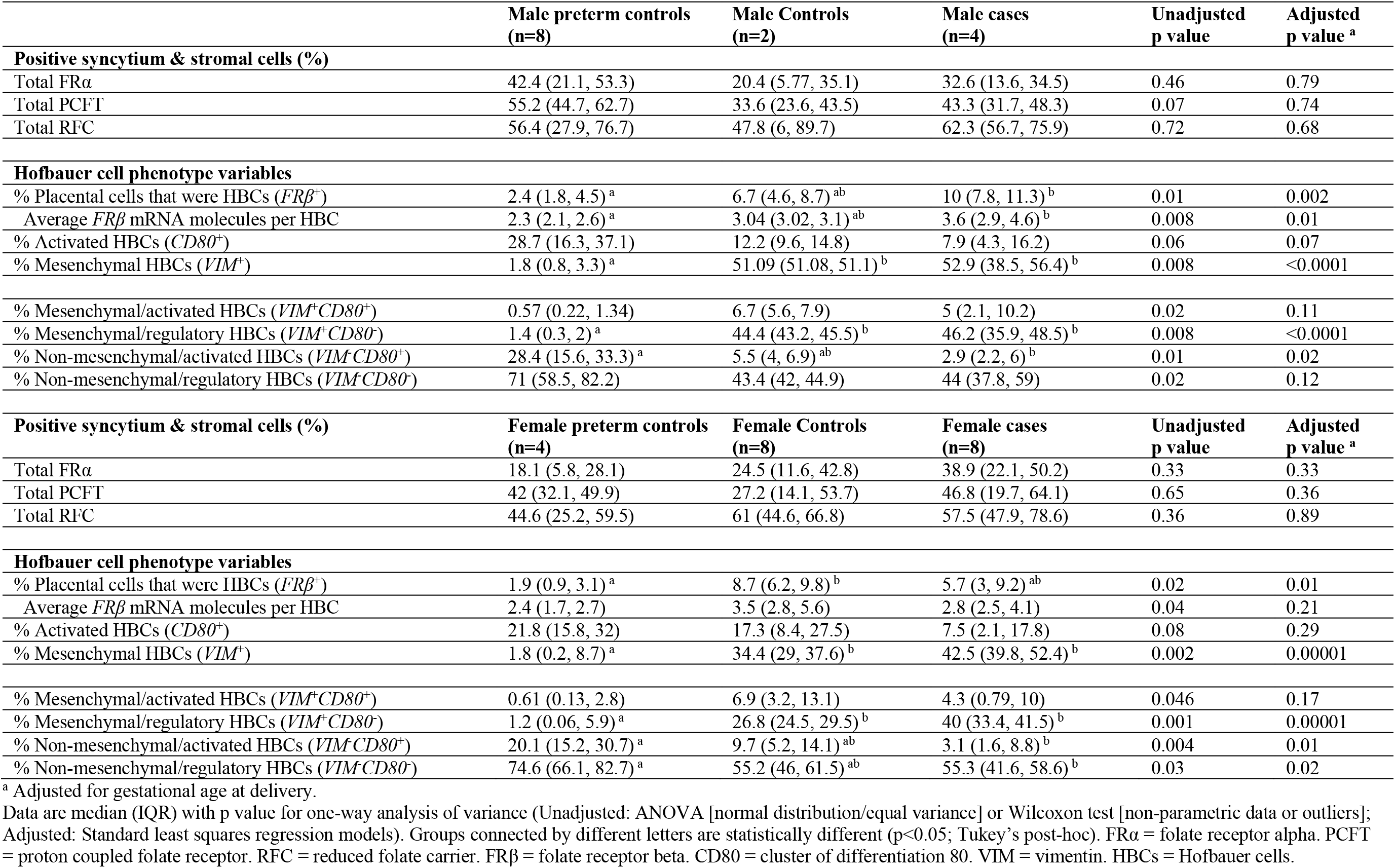
Associations between isolated spina bifida and Hofbauer cell abundance and phenotype and semi-quantitative placental folate transporter expression in male and female cases and controls.

In females, the proportion of placental villous cells that were HBCs (*FRβ*^+^ cells) was associated with study group in adjusted analyses (p=0.01; Table 1), however, *post-hoc* testing did not reveal any differences between female cases and female controls. Female cases had a higher proportion of HBCs with mesenchymal origins (*VIM*^+^) than female PT controls (42.5% [39.8, 52.4] vs. 1.8% [0.2, 8.7], p=0.00001; Table 1). There were no differences in the average *FRβ* expression by HBCs, the proportion of HBCs that were activated (*CD80*^*+*^), or FRα, PCFT, and RFC protein immunolabeling in female cases compared to female PT controls, and no differences HBC phenotype or placental folate transporter protein immunolabeling in female cases compared to female controls (Table 1).

In male compared to female cases, male cases had a higher proportion of placental villous cells that were HBCs than female cases, but the difference did not persist after adjusting for gestational age (10% [7.8, 11.3] vs. 5.7% [3, 9.2], unadjusted p=0.04, adjusted p=0.16; Table 2). There were no other differences in HBC phenotype or placental folate transporter protein immunolabeling in male cases compared to female cases (Table 2).

**Table 2.** Hofbauer cell abundance and phenotype and semi-quantitative placental folate transporter expression in male compared to female cases.

|  | Male cases (n=4) | Female cases (n=8) | Unadjusted p value | Adjusted p value <sup>a</sup> |
| --- | --- | --- | --- | --- |
| **Positive syncytium & stromal cells (%)** |  |  |  |  |
| Total FRα | 32.6 (13.6, 34.5) | 38.9 (22.1, 50.2) | 0.31 | 0.72 |
| Total PCFT | 43.3 (21.7, 48.3) | 46.8 (19.7, 64.1) | 0.73 | 0.81 |
| Total RFC | 62.3 (56.7, 75.9) | 57.5 (47.9, 78.6) | 0.5 | 0.69 |
| **Hofbauer cell phenotype variables** |  |  |  |  |
| % Placental cells that were HBCs (*FRβ*<sup>+</sup>) | 10 (7.8, 11.3) | 5.7 (3, 9.2) | 0.04 | 0.16 |
| Average *FRβ* mRNA molecules per HBC | 3.6 (2.9, 4.6) | 2.8 (2.5, 4.1) | 0.31 | 0.79 |
| % Activated HBCs (*CD80*<sup>+</sup>) | 7.9 (4.3, 16.2) | 7.5 (2.1, 17.8) | 0.87 | 0.77 |
| % Mesenchymal HBCs (*VIM*<sup>+</sup>) | 52.9 (38.5, 56.4) | 42.5 (39.8, 52.4) | 0.4 | 0.21 |
| % Mesenchymal/activated HBCs (*VIM*<sup>+</sup>*CD80*<sup>+</sup>) | 5 (2.1, 10.2) | 4.3 (0.8, 10) | 0.73 | 0.98 |
| % Mesenchymal/regulatory HBCs (*VIM*<sup>+</sup>*CD80*<sup>-</sup>) | 46.2 (35.9, 48.5) | 39.8 (33.4, 41.5) | 0.09 | 0.08 |
| % Non-mesenchymal/activated HBCs (*VIM*<sup>-</sup>*CD80*<sup>+</sup>) | 2.9 (2.2, 6) | 3.1 (1.6, 8.8) | 0.73 | 0.5 |
| % Non-mesenchymal/regulatory HBCs (*VIM*<sup>-</sup>*CD80*<sup>-</sup>) | 44.1 (37.8, 59) | 55.3 (41.6, 58.6) | 0.5 | 0.38 |

<sup>a</sup> Adjusted for gestational age at delivery.
Data are median (IQR) with p value for one-way analysis of variance (Unadjusted: Wilcoxon test; Adjusted: Standard least squares regression models). FRα = folate receptor alpha. PCFT = proton coupled folate receptor. RFC = reduced folate carrier. *FR $\beta$* = folate receptor beta. *CD80* = cluster of differentiation 80. *VIM* = vimentin. HBCs = Hofbauer cells.

### GA at delivery and maternal reported folate intake associate with placental folate transporter expression

Overall, inclusive of study group, gestational age at delivery was negatively associated with the total percent of syncytium and stromal cells that were positive for FRα protein immunolabeling (Spearman’s ρ=-0.34, p=0.049), but positively associated with the average *FRβ* expression by HBCs (ρ=0.48, p=0.004; Supplementary Figure S3).

Maternal reported folate intake (total dietary folate) was positively associated with the total percent of syncytium and stromal cells that were positive for PCFT protein immunolabeling (ρ=0.5, p=0.03; Supplementary Figure S3). There were no associations between other key maternal and infant data (maternal pre-pregnancy BMI, gestational age at delivery, birthweight z-score, and reported intakes of vitamins B6 and B12) and placental folate transporter expression or HBC phenotype in the full cohort.

Within cases, maternal reported folate intake was positively associated with the total percent of syncytium and stromal cells that were positive for RFC protein immunolabeling (ρ=0.67, p=0.03; Supplementary Figure S3). There were no associations between other key maternal and infant data and placental folate transporter expression or HBC phenotype among cases.

## Discussion

To understand whether fetal SB may associate with alterations in placental immune cell phenotype and function or placental folate transport, we have shown for the first time that *FRβ* expression by HBCs is increased in fetuses with SB, and have confirmed prior findings of increased HBC levels in these cases. We also found that a higher proportion of HBCs in cases had mesenchymal origins, but a lower proportion were of an activated *CD80*^+^ phenotype, in comparison to a control group matched to cases for GA. In exploratory sex-stratified analysis, we found that male, but not female, cases had higher HBC levels and higher average *FRβ* expression by HBCs than GA-matched controls, while female cases had a higher proportion of HBCs with mesenchymal origins than female PT controls. Folate transporter expression in the placental syncytium was not altered in fetuses with SB, in a contemporary cohort from a region where folate deficiency is rare^37^. Collectively, our findings suggest that fetal SB may associate with an increase in HBC proliferation and function, which could be a mechanism contributing to placental villous maldevelopment observed in these fetuses^12,17^ and provide further evidence in support of a role for immune dysregulation in NTDs^38^.

Our finding that cases had a higher proportion of cells that expressed *FRβ* (a marker for HBCs^14^), compared to GA-matched controls, may suggest an increase in HBC proliferation in cases. HBC proliferation has been reported in pregnancy pathologies associated with immune activation, brought about by infectious agents or immune-mediated processes^39^. As immune dysregulation is one known contributor to the pathogenesis of NTDs, and in particular, folate-resistant NTDs^38^, we hypothesised that HBC proliferation in cases may be a placental marker for an altered immune phenotype in the uterine environment. Indeed, the increase in HBC number in cases is consistent with findings from placental histopathology assessments in this cohort^17^ and a historical cohort^12^, and appears to be driven by a largely *CD80*^*-*^ HBC population, which play important regulatory roles in placental immunomodulation^34,40^. Cases also had increased *FRβ* expression by HBCs, which may be an adaptive response to a villous microenvironment that is more ‘competitive’ for folate resources, due to the high number of HBCs, or be required for an increase in HBC function. Further, we previously reported that cases in this cohort had upregulated expression of five genes known to be expressed by HBCs, including genes involved in inflammatory response regulation, which is consistent with an overall enhancement of HBC function in cases^22^. Given that HBCs regulate placental angiogenesis, and changes in HBC number or polarisation may alter angiogenesis^34^, an altered HBC phenotype may contribute to placental villous maldevelopment observed in these cases^17^. Future studies should consider a deeper phenotyping of the role of regulatory (M2) HBC subtypes (M2a, M2b, M2c, and M2d^40^) in placental physiology in both healthy fetuses, and those with isolated SB, to better delineate how HBC composition may relate to placental villous maldevelopment in fetuses with isolated NTDs.

Cases also had a higher proportion of HBCs that were of mesenchymal origins than PT controls. While HBC ontogeny across pregnancy is not well understood, as gestation progresses, fetal monocyte-derived macrophages are proposed to gradually replace the population of HBCs that arose from villous mesenchymal stem cells earlier in pregnancy^15,16^. The proportion of HBCs expressing *VIM* in PT controls, specifically, was low, which could suggest a relative increase in fetal monocyte-derived macrophages in the PT control group. It is possible that the PT controls themselves had placental dysfunction, given the role of placental maldevelopment in PT birth^8^, and notably, placental lesions have been shown to associate with fetal monocyte and monocyte subset phenotypes in fetuses born PT^41^. However, HBC polarisation was not influenced by whether HBCs were derived from mesenchymal or non-mesenchymal tissues in any of the study groups. Thus, it remains unclear whether the differences in HBC origins between study groups may relate to placental phenotype more broadly, and this is an area requiring further investigation.

We also found that male, but not female, cases had higher HBC levels and higher average *FRβ* expression by HBCs than male PT controls, which may suggest that male fetuses with SB have an increased susceptibility to immune-related alterations in placental phenotype. There are known sex differences in HBC gene expression/function and response to inflammatory stimuli^42,43^, and it is possible that altered HBC phenotypes in cases may have sex-specific effects on placental development and function^44^. While there were no differences in HBC phenotype between male and female cases after adjusting for GA, the number of male cases in this cohort was small and larger studies are needed to further explore sex-stratified and -interacting relationships between fetal SB and HBC phenotype. An improved understanding of whether placental immune function differs between male and female cases could provide insight into sex-specific NTD phenotypes and should be a focus for future research.

Because folate deficiency is a well-established risk factor for NTDs^1-3^ and may contribute to poor fetal growth^5^, we hypothesised that cases would have reduced expression of placental folate transporters, which may associate with suboptimal fetal growth. Contrary to our hypothesis, we found no differences in semi-quantitative assessments of placental folate transporter protein localisation or expression (for FRα, PCFT, and RFC) across study groups, and no associations between placental folate transporter expression and birthweight. Our findings are inconsistent with previous studies from India and Mexico, which have reported upregulated *FRα* and *RFC* expression in placentae from fetuses with NTDs ^21^, and upregulated PCFT expression in placentae from fetuses with congenital anomalies (including, but not limited to, NTDs)^45^. India does not have mandatory folic acid fortification^46^ and folic acid fortification in Mexico is not well enforced^47^, suggesting that suboptimal maternal folate status may have contributed to the changes observed in placental folate transport in these studies. However, it is likely that the SB cases in this cohort were not folate-responsive NTDs, given that folate deficiency is rare in the region the study took place^37^ and that all mothers in the cohort were taking prenatal vitamins^22^. The lack of differences in folate transporter immunolabeling reported here are consistent with transcript-level data for expression of *FRα, PCFT*, and *RFC* we have reported previously for this cohort^22^. It is possible that the suboptimal growth observed in ∼50% of the cases in this cohort was, in part, driven by dysfunction in other nutrient-related placental processes, such as lipid metabolism and amino acid transport^22^, and this is an area requiring further study. Future studies should also consider measuring maternal folate levels to investigate relationships between maternal folate status and placental folate transport in fetuses with SB.

Our use of automated analysis pipelines in QuPath to characterise placental histological features through IHC and ISH is a key strength of our study. IHC results are typically interpreted using visual semi-quantitative assessments, which can be influenced by the experience-level of the assessor and have limitations for accuracy and reproducibility^46^. Further, the use of QuPath allowed us to evaluate average marker expression over more frames of placental tissue than are typically feasible when performing assessments manually. Notably, a limitation of the QuPath analysis pipeline is that the approximated cell boundaries may not exactly reflect true cell boundaries. Despite that cell boundaries were estimated based on known HBC size and probes could only be counted once, it is possible this approximation could have led to the misclassification of target probe co-expression for HBCs. Additionally, although advantages of RNA *in situ* hybridization include that the method is highly specific and quantifiable, to the level of a single molecule, and we used three successful markers to identify and characterise HBC populations, HBCs are highly heterogeneous and a deeper characterisation of HBC subtypes (e.g., M2a, M2b, M2c, and M2d, each of which differ in function, surface marker expression, and cytokine secretions^38^) using additional markers and methods could improve our understanding of how HBC phenotype is altered in fetuses with isolated NTDs. Further, while *FRβ* has been shown to be a highly specific marker for identifying HBCs^47^, *FRβ* is also expressed by other myeloid cells that may be present in the placental villous stroma, such as monocytes^48^. Notably, increasingly throughout pregnancy and in term placenta, HBCs are proposed to be monocyte-derived^15,16^, suggesting that if any monocytes were identified, they may have been precursors to HBCs. Finally, as our sample size was limited to 12 cases, replicating these findings in other cohorts to understand their validity and generalizability should be a focus of future study.

This study is the first to characterise HBC phenotypes in fetuses with SB, and the first to evaluate placental folate transport in a cohort of fetuses with isolated SB in Canada. Understanding how HBC phenotypes are altered in fetuses with SB helps to increase knowledge on the immune-related factors that may contribute to NTD pathogenesis, phenotype, and NTD-associated comorbidities^38^. That placental folate transport in cases did not differ from fetuses without congenital anomalies is promising, but affirms the need for ongoing research to better delineate the folate-independent mechanisms that continue to drive fetal NTDs and associated comorbidities.

## Data Availability

The data that support the findings of this study are available on reasonable request from the corresponding author, KLC.

## Acknowledgements

The authors would like to thank Vagisha Pruthi for overseeing the study recruitment, data entry and sample collection, the staff at the Research Centre for Women’s and Infants’ Health (RCWIH) BioBank for collecting and processing the placental samples, and the staff at the Louise Pelletier Histology Core Facility and the Cell Biology and Image Acquisition Core Facility for their assistance with slide imaging.

This research is supported by the Canadian Institutes of Health Research (CIHR PJT-175161) and departmental research grants from the Department of Obstetrics and Gynaecology at Mount Sinai Hospital, Toronto, and the Department of Health Sciences at Carleton University, Ottawa. MW was supported by an Ontario Graduate Scholarship, Faculty of Science, Carleton University.

## Author contributions

Conceptualization: MW, KLC, TVM, and DG; Resources: TVM and KLC; Investigation: MW, KLC, HA, DG; Data curation: MW and HA; Methodology: MW, KLC, HA; Formal analysis: MW and HA; Visualization: MW; Writing – original draft preparation: MW and KLC; Writing – reviewing & editing: MW, KLC, TVM, HA, and DG. Funding acquisition: KLC and TVM; Supervision: KLC.

