## Supplementary Figures for "Altered placental immune cell composition and gene expression with isolated fetal spina bifida"

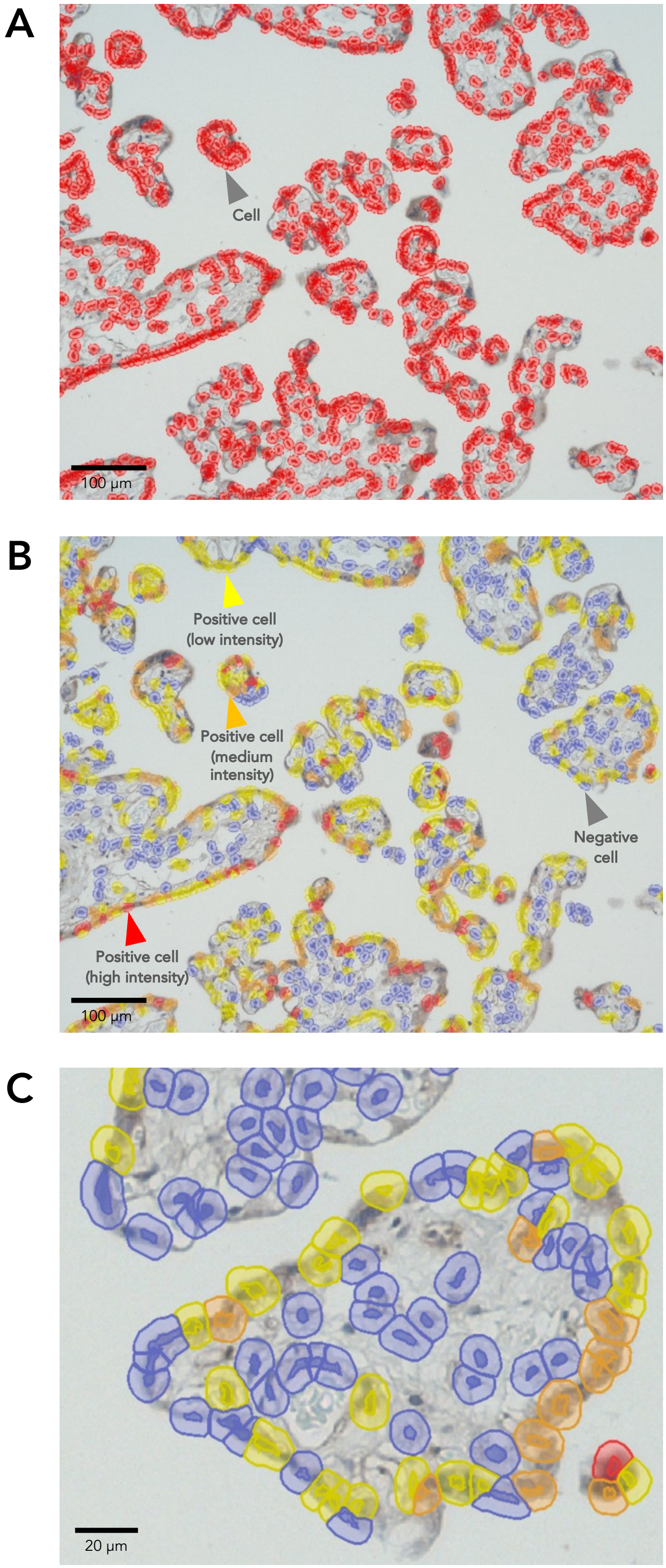


**Supplementary Figure S1. Example images of the detection methods applied to semi-quantitatively evaluate protein staining of immunolabeled areas in placental syncytium and stromal cells.** (A) View of a placental field (image) containing placental villous tissue annotated after cell detection parameters were applied. Outlines for the cell nucleus and predicted cell boundary (5 µm cell expansion) are shown in red. (B-C) Example views of placental tissue after positive cell detection parameters were applied. Cells were classified as negative for protein staining (blue), or positive with low (yellow), medium (orange), or high (red) intensity staining. Panel C is a closer view of a section of the image in panel B. The example images are from a placental sample treated with PCFT antibody.

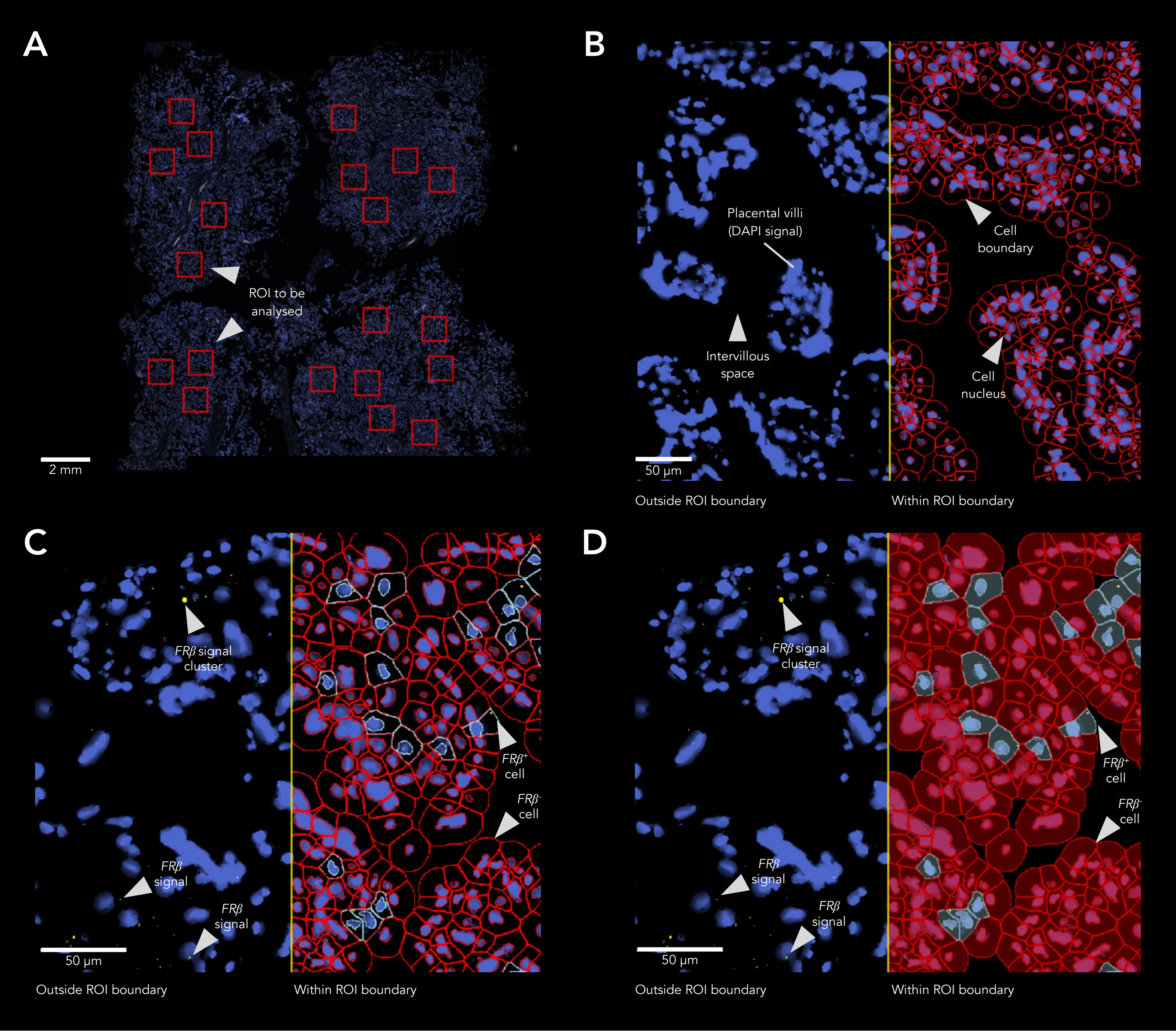


**Supplementary Figure S2. Example images of cell and probe detection methods applied to characterize Hofbauer cell phenotypes in fluorescent images in QuPath.** (A) Whole-slide scan view of a placental sample with 20 regions of interest (ROI; 1000 µm x 1000 µm squares) containing placental villous tissue annotated for analysis. ROI annotations (red squares) were determined using only the DAPI signal (with all other detections hidden), as is represented here. (B) Example view of placental tissue after cell detection parameters were applied. Outlines for the cell nucleus and predicted cell boundary (10 µm cell expansion) are shown. (C-D) Example views of placental tissue after probe detection thresholds were applied. Cells were tagged as probe$^{+}$ (in the example frame [C-D], *FRβ*$^{+}$; cells marked in turquoise) if *FRβ* signal (one or more punctate dots) was detected within the cell boundary. Red cells were *FRβ*$^{-}$. Panels C and D show the same frame, but with different detection viewing settings to more easily visualize probe localization (C) and *FRβ*$^{+}$ vs. *FRβ*$^{-}$ cell distribution. In panels B-D, the left side of the panel shows placental tissue that was not annotated (outside of the ROI boundary), and the right side of the panel shows annotated tissue (within the ROI boundary).

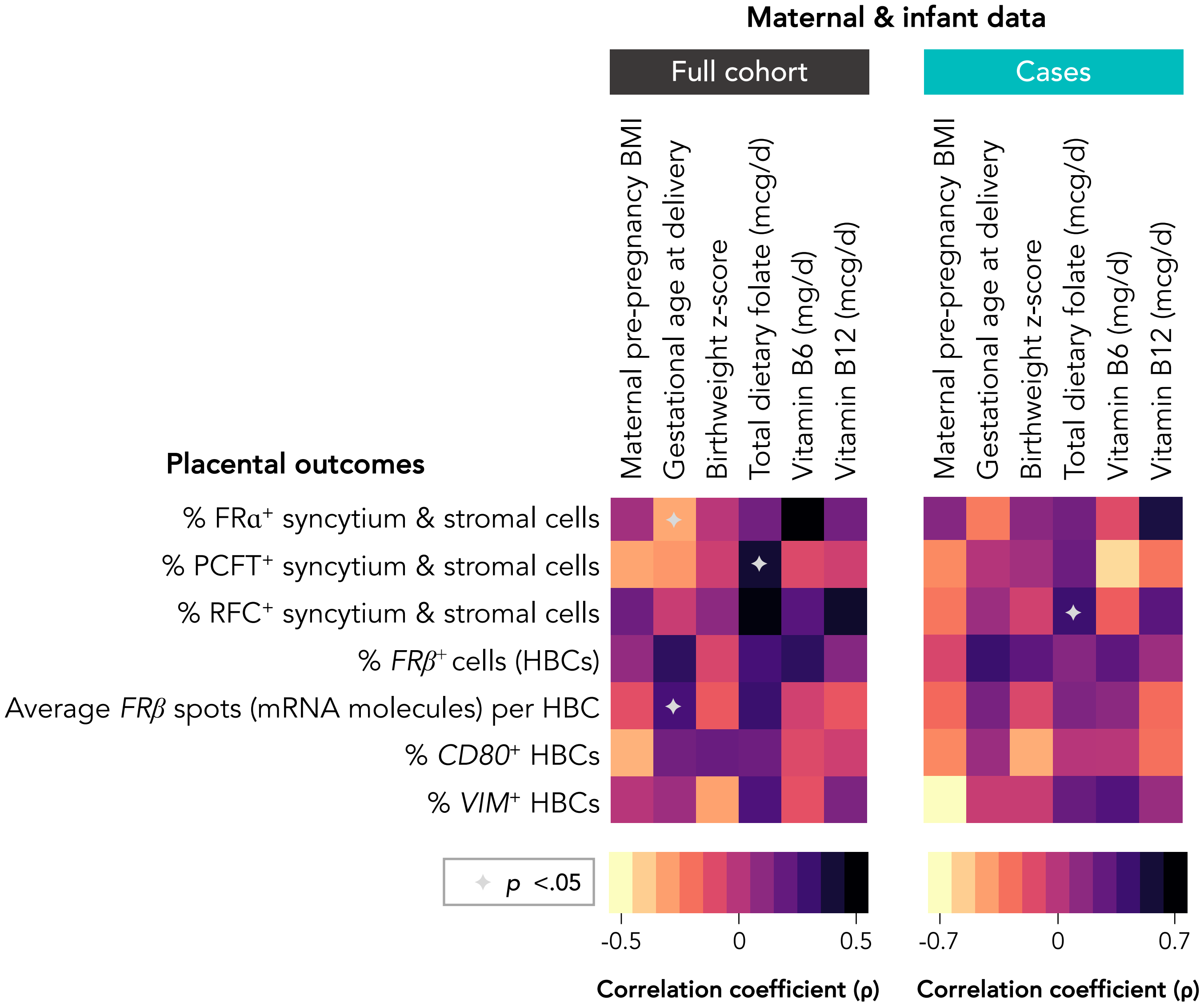


**Supplementary Figure S3. Associations between key maternal and infant data, and folate-sensitive placental outcome variables.** Data are from Spearman's rank correlation (rho; **ρ**) test with p values. Spearman's **ρ** are represented in heatmaps (yellow-to-orange colours=negative correlation, pink-to-black colours=positive correlation). Statistical significance is denoted by grey symbols (✦ = p value <0.05). Full cohort = all samples inclusive of study group.
