## Supplementary Tables for "Altered placental immune cell composition and gene expression with isolated fetal spina bifida"

**Supplementary Table S1.** Positive cell detection settings used in QuPath for semi-quantification of immunoreactive FRα, PCFT and RFC staining.

| | |
|---|---|
| **Setup parameters** | |
| Detection image | Optical density sum |
| Requested pixel size | 0.2 µm |
| **Nucleus parameters** | |
| Background radius | 8 µm |
| Median filter radius | 1 µm |
| Sigma | 0 µm |
| Minimum area | 10 $\mu m^2$ |
| Maximum area | 400 $\mu m^2$ |
| **Intensity parameters** | |
| Threshold | 0.25 |
| Max background intensity | 2 |
| Split by shape | Yes |
| Exclude DAB (membrane staining) | No |
| **Cell parameters** | |
| Cell expansion | 5 µm |
| Include cell nucleus | Yes |
| **General parameters** | |
| Smooth boundaries | Yes |
| Make measurements | Yes |
| **Intensity threshold parameters** | |
| Score compartment | Cell: DAB OD mean |
| Threshold 1+ | 0.15 |
| Threshold 2+ | 0.20 |
| Threshold 3+ | 0.25 |
| Single threshold | No |

**Supplementary Table S2.** Cell detection settings used in QuPath to detect DAPI signal and probe detection thresholds for single measurement object classifiers.

| | |
|---|---|
| **Setup parameters** | |
| Detection channel | DAPI |
| Requested pixel size | 0.4 µm |
| **Nucleus parameters** | |
| Background radius | 8 µm |
| Median filter radius | 0 µm |
| Sigma | 1.5 µm |
| Minimum area | 10 $\mu m^2$ |
| Maximum area | 400 $\mu m^2$ |
| **Intensity parameters** | |
| Threshold | 40 |
| Split by shape | Yes |
| **Cell parameters** | |
| Cell expansion | 10 µm |
| Include cell nucleus | Yes |
| **General parameters** | |
| Smooth boundaries | Yes |
| Make measurements | Yes |
| **Probe detection thresholds for single measurement object classifiers** | |
| *FRβ* | 1 (Cell: Opal 620 mean) |
| *CD80* | 0.35 (Cell: Opal 690 mean) |
| *VIM* | 150 (Cell: Opal 570 mean) |

**Supplementary Table S3.** Associations between isolated spina bifida and semi-quantitative placental folate transporter expression in cases and controls.

| | Preterm controls (n=12) | Controls (n=10) | Cases (n=12) | Unadjusted p value | Adjusted p value † |
|---|---|---|---|---|---|
| **Positive syncytium & stromal cells (%)** | | | | | |
| Total FRα | 27.8 (13.6, 50.1) | 24.5 (8.4, 40.5) | 34.4 (22.1, 41.3) | 0.8 | 0.51 |
| FRα low intensity threshold | 18.1 (11.2, 27.9) | 14.3 (5.6, 21.5) | 19.3 (12.4, 20.6) | 0.53 | 0.56 |
| FRα medium intensity threshold | 7.2 (2, 14.4) | 5.9 (1.8, 10.3) | 8.1 (5.4, 11.2) | 0.8 | 0.49 |
| FRα high intensity threshold | 2.7 (0.37, 6.7) | 4.7 (1.1, 7.7) | 6.4 (3.4, 9.6) | 0.16 | 0.4 |
| Total PCFT | 50.6 (41.7, 61) | 28.7 (16.8, 52.7) | 44.6 (24.6, 51.9) | 0.13 | 0.63 |
| PCFT low intensity threshold | 21.6 (21.1, 26.8) | 16.3 (9.8, 22.5) | 19.5 (13.4, 23.4) | 0.02 | 0.21 |
| PCFT medium intensity threshold | 14.6 (11.8, 17.1) | 7.1 (3.9, 14.6) | 12.1 (6.1, 14.7) | 0.07 | 0.41 |
| PCFT high intensity threshold | 10.6 (6.7, 16.4) | 6.2 (2.2, 13.4) | 11.1 (5.4, 13.2) | 0.33 | 0.97 |
| Total RFC | 53.5 (27.9, 62) | 61 (41.6, 68.2) | 59.6 (55.1, 75.9) | 0.48 | 0.45 |
| RFC low intensity threshold | 17.2 (15.8, 23) | 23.1 (17.5, 24.7) | 21.8 (16.9, 25.1) | 0.34 | 0.64 |
| RFC medium intensity threshold | 13.6 (7.2, 18.3) | 14.1 (11.2, 18.4) | 16.5 (14.8, 18.2) | 0.66 | 0.53 |
| RFC high intensity threshold | 14.8 (4.5, 23.5) | 22.2 (7.3, 25.7) | 18.7 (17.3, 36.5) | 0.53 | 0.47 |

† Adjusted for infant sex and gestational age at delivery.
Data are median (IQR) with p value for one-way analysis of variance (Unadjusted: ANOVA [normal distribution/equal variance] or Wilcoxon test [non-parametric data or outliers]; Adjusted: Multiple linear regression models). Groups connected by different letters are statistically different ($p<0.05$; Tukey's post-hoc). FRα = folate receptor alpha. PCFT = proton coupled folate receptor. RFC = reduced folate carrier.

**Supplementary Table S4.** Associations between isolated spina bifida and placental Hofbauer cell abundance and phenotype in cases and controls.

| | **Preterm controls (n=12)** | **Controls (n=10)** | **Cases (n=12)** | **Unadjusted p value** | **Adjusted p value** † |
|---|---|---|---|---|---|
| **Hofbauer cell phenotype variables** | | | | | |
| % Placental cells that were HBCs (*FRβ*$^+$) | 2.4 (1.2, 3.2) [a] | 8.6 (5.7, 9.5) [b] | 6.9 (4.3, 10.2) [b] | 0.0002 | 0.0001 |
| Average *FRβ* mRNA molecules per HBC | 2.3 (2.1, 2.6) [a] | 3.1 (2.9, 4.5) [ab] | 3.2 (2.5, 4.2) [b] | 0.0003 | 0.03 |
| % Activated HBCs (*CD80*$^+$) | 26.6 (15.8, 33.3) [a] | 13.8 (8.9, 23.8) [ab] | 7.9 (2.6, 17.7) [b] | 0.003 | 0.01 |
| % Mesenchymal HBCs (*VIM*$^+$) | 1.8 (0.7, 3.3) [a] | 36 (31.4, 43.7) [b] | 43.7 (39.8, 54.9) [b] | <0.0001 | <0.0001 |
| % Mesenchymal/activated HBCs (*VIM*$^+$*CD80*$^+$) | 0.61 (0.22, 1.3) [a] | 6.7 (3.5, 11.2) [b] | 5 (0.9, 10) [b] | 0.0005 | 0.02 |
| % Mesenchymal/regulatory HBCs (*VIM*$^+$*CD80*$^-$) | 1.4 (0.24, 2.1) [a] | 28.4 (25.3, 36.8) [b] | 40.3 (33.4, 45.4) [b] | <0.0001 | <0.0001 |
| % Non-mesenchymal/activated HBCs (*VIM*$^-$*CD80*$^+$) | 24.8 (15.2, 32.5) [a] | 7.3 (4.9, 13.5) [b] | 3 (1.9, 6.3) [b] | <0.0001 | 0.00003 |
| % Non-mesenchymal/regulatory HBCs (*VIM*$^-$*CD80*$^-$) | 71 (66.1, 82.7) [a] | 51.5 (44.3, 60) [b] | 50.1 (41.1, 58.6) [b] | 0.001 | 0.0002 |
| % Of mesenchymal HBCs that were activated (*CD80*$^+$) | 40 (31.1, 52.6) [a] | 15.2 (10.5, 29.5) [b] | 9.4 (2.4, 19) [b] | 0.001 | 0.0002 |
| % Of non-mesenchymal HBCs that were activated (*CD80*$^+$) | 26.2 (15.6, 33) [a] | 12.9 (7.6, 21.2) [ab] | 6.3 (2.8, 14.5) [b] | 0.001 | 0.003 |

† Adjusted for infant sex and gestational age at delivery.

Data are median (IQR) with p value for one-way analysis of variance (Unadjusted: ANOVA [normal distribution/equal variance] or Wilcoxon test [non-parametric data or outliers]; Adjusted: Multiple linear regression models). Groups connected by different letters are statistically different ($p<0.05$; Tukey's post-hoc). *FRβ* = folate receptor beta. *CD80* = cluster of differentiation 80. *VIM* = vimentin. HBCs = Hofbauer cells.
